# Assessing Legal Protections for Care Workers: A 10-Country Analysis

**DOI:** 10.64898/2026.03.31.26349840

**Authors:** Vishakh Unnikrishnan, Eric Friedman, Matthew M. Kavanagh, Catherine Kane

## Abstract

Care workers are central to health systems and the broader care economy, but they often lack the legal protections afforded to other workers. Furthermore, there currently exists no single legal definition of "care worker" under any binding or non-binding international legal instrument. Drawing on the WHO Global Health and Care Worker Compact, we analyzed whether national laws and policies in 10 countries protect care workers. Using comparative legal methods and primary source legal and policy documents, we evaluated both care worker coverage and alignment with four indicators: guaranteed access to protective equipment, protection against discrimination on internationally recognized grounds, unemployment insurance, and the right to join independent unions. We reviewed 43 laws and policies and found that 56% fully or partially met the relevant indicator criteria. The United Kingdom was the only country meeting all four indicators. Overall, we found while many countries recognize these protections in law, care workers are often left outside their coverage, underscoring the need for clearer legal recognition and more inclusive worker protections.

## Introduction

Care workers globally, including those working in institutional and residential settings, and whether they work in the formal or informal sector, often lack the legal protections afforded to other workers. Global estimates suggest only 10% of domestic workers enjoy labor protections equivalent to those enjoyed by other workers and over 90% of domestic workers are legally excluded from social security systems [1]. Care work is often undervalued, with many care workers citing systemic issues as a reason, including race- and gender-based discrimination and xenophobia [2, 3]. Care workers also face challenges working in unregulated work environments such as private households [1]. In addition, they may have multiple employers, often without a formal employment contract while working in a precarious or hazardous work environments.. They include those who work in provide personal care in the health, social work, education, and domestic work sectors, among others [4].

Care workers, particularly those working in domestic settings, face heightened risks to their health, safety, and rights. A Human Rights Watch report found migrant domestic workers routinely encounter exploitative working conditions, including excessively long working hours, lack of rest days or rest periods, poor living accommodations, and restrictions on freedom of movement and association [5]. Migrant domestic workers also face challenges involving their immigration status and challenges related to their right to work in their country of residence, which may, for example, impede their ability to receive social security and other benefits [5, 6]. Furthermore, 14% of countries that provide some social security coverage for domestic workers do not provide the same rights to migrant domestic workers [1].

Notably, even as formal care workers frequently lack protections afforded to other workers, much care work goes unpaid altogether. A 2018 report states over 16.4 billion hours are spent in unpaid care work every day – equivalent to 2 billion people working eight hours per day with no remuneration [4]. Unpaid care work represent approximately 9% of global GDP, equivalent to $11 trillion, highlighting unpaid care workers’ economic contribution despite being uncompensated [7]. Women make up 67% of the global health and social care workforce and perform 76% of unpaid care activities [8].

Care, both paid and unpaid, is fundamental to well-being. A strong and effective care economy promotes a healthier current and future workforce, generates employment opportunities, supports businesses, and boosts overall productivity. The COVID-19 pandemic highlighted the critical role of the care economy and amplified the unequal distribution of unpaid care work, with these responsibilities falling primarily to women and girls. It underscored the longstanding issue of unsafe working conditions faced by care workers including long working hours, occupational health and safety risks, and exposure to violence and harassment, including gender-based violence [9].

Recognizing the challenges and difficulties health and care workers face, especially in the midst of the COVID-19 pandemic, the 74th World Health Assembly [WHA] in 2021 requested the WHO Director-General to develop a global health and care worker compact [hereafter referred to as the "Care Compact"] [10]. The Care Compact, a technical compilation of international labor and human rights legal instruments, including treaties and existing normative documents of relevant international organizations such as the World Health Organization [WHO] and International Labor Organization [ILO], provides guidance on “protecting health and care workers and safeguard their rights, and to promote and ensure decent work, free from racial and all other forms of discrimination and a safe and enabling practice environment ”[10]. The following year, with the Care Compact complete, the WHA encouraged states to use their Care Compact to guide their actions to “protect and support health and care workers,” and for the Director-General to utilize the Care Compact in supporting states to safeguard health and care workers’ rights and fair and decent working conditions [11].

The Care Compact is thus a central document for countries to inform and implement measures to ensure the protection of rights of both health and care workers. It categorizes legal and policy instruments across four domains comprising 10 areas that are linked to human and labor rights. The four domains are preventing harm, inclusivity, providing support, and safeguarding rights. These domains focus on ensuring the protection of health and care workers, enabling their rights and protections, providing financial and non-financial support, and empowering collective and individual rights [10].

For our analysis, we chose four indicators, one in each of these four domains. *Table 1* provides a description of each of these domains while *Table 2* provides our rationale for choosing these benchmarks for each indicator.

**Table 1:** Care Compact Indicator Descriptions.

| Care Compact Domain | Description |
| --- | --- |
| Right to Safe and Healthy Working Conditions | Article 7 of the ICESCR ensures fair wages, safe and healthy working conditions, and equal opportunity in employment for all workers, *de facto* including all care workers [12]. ILO conventions such as the |
|  | Occupational Safety and Health Convention, 1981 (No. 155) and the Promotional Framework for Occupational Safety and Health Convention, 2006 (No. 187) likewise apply to all workers [13, 14]. ILO's Resolution on the inclusion of a safe and healthy working environment also obligates all member states to ensure promote and to realize the principles concerning the fundamental right to a safe and healthy working environment [15]. These international legal instruments emphasize the need for occupational safety, place the onus on employers to ensure worker safety, and require provision of essential protective equipment for all workers. |
| **Non-Discrimination** | Human rights treaties, including the Convention on the Elimination of All Forms of Discrimination Against Women (CEDAW) and the Convention on the Rights of Persons with Disabilities (CRPD), mandate non-discrimination and equal treatment in the workplace [16, 17]. The ILO's Equal Remuneration Convention, 1951 (No. 100), Social Security (Minimum Standards) Convention, 1952 (No. 102) and Discrimination (Employment and Occupation) Convention, 1958 (No. 111) similarly emphasize fair wages and social protection for all workers and protection against discrimination at the workplace [18-20]. These laws clearly outline the rights and protections applying to all workers. |
| **Social protection** | The ILO's Social Security [Minimum Standards] Convention, 1952 (No. 102) and Unemployment Promotion and Protection against Unemployment Convention, 1988 (No.168), and the ICESCR1966, sets minimum standards and emphasizes unemployment protection for all workers, as part of social protection. Other non-binding agreements such as the Recommendation on Social Protection Floors, 2012 (No. 202) and Global Jobs Pact, 2009 encourage member states to promote policies that offer social protection to all workers without distinguishing between formal and informal workers [18, 21-23]. |
| **Right to Collective Bargaining** | Binding international instruments, such as the Freedom of Association and Protection of the Right to Organize Convention, 1948 (No. 87), the Right to Organize and Collective Bargaining Convention, 1949 (No. 98), and the and International Covenant on Civil and Political Rights (ICCPR), 1966, establish the fundamental right of workers and employers to form and join a union independent of government or employer interference [24-27]. The Universal Declaration on Human Rights, now binding customary international law, guarantees the right to form and join unions, as well as the right to freedom of association. Other, non-binding, instruments, such as the Declaration on Fundamental Principles and Rights at Work, 1998 and the Global Jobs Pact, 2009, encourage member states to promote and respect the right to freedom of association and the right to collective bargaining [28-30]. |

International instruments, encompassing both binding and non-binding agreements—such as the International Covenant on Economic, Social and Cultural Rights [ICESCR] and International Labour Organization [ILO] conventions and recommendations—underscore the right to “decent work”: safe and healthy working conditions for all workers, making no distinction between formal workers and informal workers, or between institutional and residential settings. These treaties and other instruments thus confer comparable rights and protections upon care workers as those afforded to all other workers.

Recognizing that the protections and rights outlined above apply universally to all workers, we examine the extent to which rights and protections are in fact afforded to care workers. Foundational instruments outlined in the Care Compact, both binding and non-binding, assert universal access to safe workplaces and obligate employers to protect care workers. Additional protections against workplace discrimination, enshrined in human right treaties and ILO conventions, as outlined in *Table 1,* reinforce this coverage for care workers, while also further extending minimum safety nets and unemployment benefits to all workers, underscoring a stated political commitment to equality [16-18, 21]. By integrating these principles, the Care Compact positions care work as labor equally deserving of robust protections, advancing a universalist approach that elevates the rights and security of those working in this vital sector.

A previous study analyzing law and policies environments that offer protection to health workers in 182 countries found that 62% of all national laws and policies aligned with all ten indicators assessed. Many of the indicators did also address care workers, but by assessing whether both health and care workers were covered, rather than assessing care workers separately [31]. We look separately at coverage of care workers in our study partly because care workers are often not covered under laws and policies that apply to other occupations including health workers, and at times have exclusive policies and laws that focus on a sub-group of care workers such as domestic workers [32]. The objective of this paper was to examine whether countries address care workers in national laws and policies across various aspects of the Care Compact [31].

### Defining Care Workers

There currently exists no single legal definition of "care worker" under any binding or non-binding international legal instrument. However, the ILO defines care work more broadly as encompassing a range of tasks that address the physical, psychological, and emotional needs of individuals across the life course, including both direct and indirect care activities aimed at sustaining and enhancing quality of life [9]. By this definition, care workers include and are often dominated by women and migrant workers. In all socioeconomic contexts, women perform three-quarters of unpaid care work across the world [4, 8]. In 2022, migrant women were nearly twice as likely as non-migrant women to be employed in care work, highlighting the extent to which care work is shaped by women and migrant labor, and underscoring the need for clearer definitional and legal recognition of who is considered a care worker [33]. This was also reflected during the COVID-19 pandemic when care workers, who often come from marginalized groups, were among the most exposed to occupational hazards, and were often at the forefront of providing direct health care services, contributing to high rates of infection and death. Yet even with their heightened risk, they tended to have limited social protections [34]. While unpaid care within private household is overwhelmingly shouldered by women and girls, it is generally excluded from the scope of “work” in labor and social protection law—because it lacks a formal employment relationship, contract, or agreed remuneration—and is therefore not included in our analysis of care workers.

## Methods

For our analysis, we adopted the Care Compact’s approach of defining care workers based on the International Labour Organization International Standard Classification of Occupations (ISCO-08) [10, 35]. This includes two occupational groups: 1) institution-based personal care workers who provide direct personal care and assistance with activities of daily living to patients and residents in various health care settings, and 2) home-based personal care workers who provide routine personal care and assistance with Activities of Daily Living] in private homes and other independent residential settings. To determine whether care workers employed in both institution- and home-based settings, as per the definitions adopted by the Care Compact, were covered under national level legislation and laws, we adopted a multi-pronged approach. This included examining explicit mentions of “informal” or “domestic” workers or other parallel classifications, evaluating whether definitions of employer-employee relationships encompassed the informal sector, and determining whether the relevant laws applied universally to all individuals employed across various workplaces. While domestic workers are not explicitly referenced in the Care Compact, domestic workers can fall under the definition of home-based care workers, as they provide personal care in private settings [36].

### Developing Indicators

To assess the alignment of laws and policies with our indicators, our first step was to develop measurable benchmarks for each as described below:

- For occupational health, we used legally guaranteed access to personal protective equipment (PPE).
- For equal treatment and non-discrimination, we used presence of policies covering internationally protected grounds set forth by the Committee on Economic, Social and Cultural Rights, which monitors implementation of the International Convention on Economic, Social and Cultural Rights.
- For social protection, we used unemployment insurance coverage.
- For collective bargaining, we used labor standards on unionization prescribed by the International Labor Organization [37, 38].

*Table 2* provides our rationale for choosing these benchmarks for each indicator.

**Table 2:** Indicator Questions and Coding Rules for Assessing Alignment of National Laws and Policies.

| *Indicator* | *Indicator Criteria* | *Rationale* |
| --- | --- | --- |
| **1. Protection from occupational hazards** | Legally guaranteed access to protective equipment or supplies | PPE has been shown to prevent the spread of disease and preserve the health of individual practitioners including care workers. This has especially been shown to be effective during the COVID-19 pandemic [39]. Furthermore, ILO's Occupational Safety and Health Convention mandates employers to provide protective equipment at the workplace [40]. General Comment No. 23 (2016) of the Committee on Economic, Social and Cultural Rights underscores the obligation of states to ensure safe and healthy working environments as a fundamental aspect of the right to just and favorable conditions of work [41]. |
| **2. Equal treatment and non-discrimination** | Prohibits discrimination on internationally protected grounds | This is based on the Committee on Economic, Social and Cultural Rights' General Comment No. 20 on non-discrimination protections. Adopted in 2009, this general comment provides a detailed framework on the obligation of states to prevent discrimination and promote equality in the enjoyment of economic, social, and cultural rights [38]. This is the most comprehensive, authoritative interpretation of internationally prohibited grounds of discrimination. |
| **3. Social protection** | Unemployment Insurance Coverage | Globally, unemployment insurance schemes have been shown to help to prevent workers and their families from falling into poverty during unemployment periods and stabilize household income [42]. |
| **4. Freedom of association and collective bargaining** | Right to Unionize and Independence. | We followed the labor standards set forth by the *Freedom of Association and Protection of the Right to Organise Convention, 1948 (No. 87)* as established by the International Labour Organization (ILO) [37], one of ILO's "fundamental instruments" [43]. Also, the CCPR explicitly states the right to freedom of association with others, including the right to form and join trade unions as a basic human right essential for the protection of individual and collective interests [44]. |

### Selection of Countries

Our country selection process for this analysis followed a two-step approach: first, ensuring representation from each of the WHO’s six designated regions, and second, confirming the availability of publicly accessible primary source documents, such as laws or policies, for analysis. We also considered whether primary source documents were originally in English or a version translated into English was available. While some of the countries selected may have particular geopolitical and economic influence in their regions, this was not a criterion for selection. However, the wide range of countries does help provide examples of care worker protection measures, or lack thereof, in various regional settings. The chosen countries are Canada [Region of Americas], Germany [European Region], Turkey [European Region], Argentina [Region of Americas], Malaysia [Western Pacific Region], United Kingdom [European Region], Angola [African Region], Ethiopia [African Region], Bangladesh [South East Asian Region], and Morocco [Eastern Mediterranean Region].

### Analyzing laws and policies

We relied solely on primary source law and policy documents to determine the extent to which the legal and policy environments in our selected countries conformed to our indicators. The process of evaluating alignment to an indicator involved two steps: first, determining whether the applicable law and policy covered all care workers and, second, whether it aligned with our indicator question. These two steps were conducted independently. A law or policy document was marked as “aligned” if our analysis showed it covered care workers and met the criteria of the relevant indicator question. If a law and policy did not cover care workers, it was marked “not aligned” regardless of whether or not it met the indicator criteria. For three of the indicators – those on occupational health, prevention of discrimination, and the right to join a union –certain laws and policies were categorized as partially aligned if they ensured care worker coverage and met some, but not all, of the criteria of the indicator question. *Table 2* provides more details about the coding rule developed to translate information about law and policies into a measure of alignment with our indicators.

**Table 3:** Indicator Questions and Coding Rules Used to Assess Whether National Laws and Policies Cover and Protect Care Workers.

| Indicator | Indicator Question | Coding Rule |
| --- | --- | --- |
| **1. Protection from occupational hazards** | Does the country have national policies or laws for occupational health and safety that cover care workers and require them to be ensured access to protective equipment or supplies? | Occupational hazard laws/policies that explicitly guarantee access to personal protective equipment to care workers are aligned (Y). Those that cover care workers but do not mandate employees to provide PPE, or where PPE provision requirements are limited to certain settings, are coded partial (P). Those that do not cover care workers are not aligned (N) regardless of PPE status. |
| **2. Equal treatment and non-discrimination** | Does the country have a law that prohibits discrimination in the workplace, on internationally protected grounds, that includes care workers? | Laws/policies that cover care workers and prohibit workplace discrimination on all internationally recognized grounds are aligned (Y), while laws that cover only some grounds are partial (P). Laws that do not cover care workers are not aligned (N) regardless of inclusion of internationally protected grounds. |
| **3. Social protection** | Does the country have national social protection policies or laws that, at minimum, provide unemployment insurance to all care workers? | Social protection laws that include care workers and provide at minimum unemployment insurance to care workers are aligned (Y). This can include the option of opting-in where care workers are permitted to avail unemployment insurance if they wish to do so. |
| **4. Freedom of association and collective bargaining** | Does the country have national policies or laws that protect the right of care workers to join an independent union or similar worker's organization? | Laws and policies that grant care workers the right to join an independent union are aligned (Y). Laws that guarantee the right to join a union, but do not ensure the union's independence, are classified as partial (P). Laws that do not include care workers are not aligned (N). Independence is assessed per international labor norms on union registration, formation, and membership independence from government [37]. |

Laws and policies were assessed using comparative legal research methodology with a focus on both descriptive comparisons.[45] This involved analyzing the content of the law and evaluating their effectiveness in aligning with our indicators. To determine care worker coverage, we sought to interpret the intent of the law or policy rather than a strict literal reading of the text [46]. Coverage of care workers was determined through several interpretative methods consistent with this approach, including broad interpretations of “employer-employee” relationship, characterized by defining employee as anyone with a working relationship with an employer or anyone with a contract of service with an individual or an organization, and employment contracts that involve provision of personal services, both of which were interpreted as inclusive of care workers. Laws and policies that explicitly mention “informal workers” or “domestic workers,” or other parallel classifications, were also understood to cover care workers within their scope. In instances where constitutional provisions were analyzed to address indicator questions, it was presumed that care workers were covered under the broader protections provided under the constitution. This presumption was based on the general application of constitutional rights and guarantees to all workers, absent explicit exclusions. Country-specific legal analysis of every law and policy in this paper is available in *Annex 1*.

## Results

Across all 4 indicators in 10 countries, we analyzed 43 different laws and policies and found 56% (24 in total) of them to have fully or partially met the indicator criteria it was being evaluated against. Of countries analyzed, only the United Kingdom’s national laws and policies met our criteria for all four indicators. All countries fully or partially met at least one of the four indicator criteria. Angola, Argentina, and Germany had national policies aligned with three of the four indicators, whereas Canada and Turkey did not meet three of the four indicator criteria.

Across the indicators on non-discrimination protections, social protection, and collective bargaining we found several countries to have laws or policies that completely or partially met our specific indicator criteria but did not include care workers within their scope of coverage (*Table 3*). For example, Canada has laws and prohibit workplace discrimination on all internationally recognized grounds; however, they only apply to formal health workers and are restricted to specific sectors. Similarly, for indicators on occupational health, non-discrimination, and social protection, we found examples where national laws and policies for countries ensured coverage of care workers but did not meet our specific indicator criteria. For example, Bangladesh’s national social protection law ensures care worker coverage but only provides severance pay without any provision of unemployment insurance. *Figures Y* and *Figure Z* present the distribution of indicator codes across all countries reviewed, distinguishing between laws and policies that cover care workers and those that do not.

**Figure Y:**
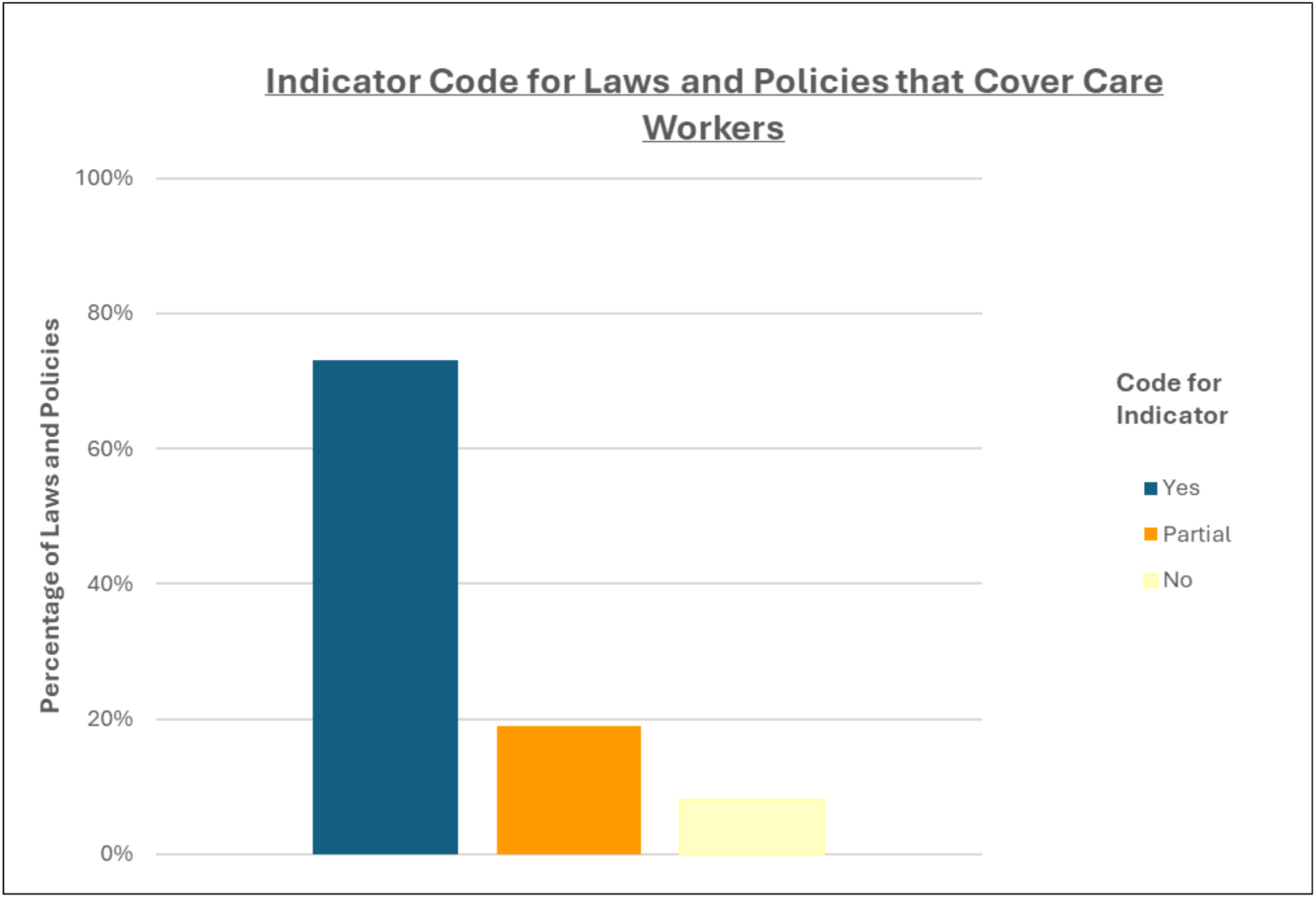
Alignment of Laws and Policies Covering Care Workers With Indicator Criteria.

**Figure Z.**
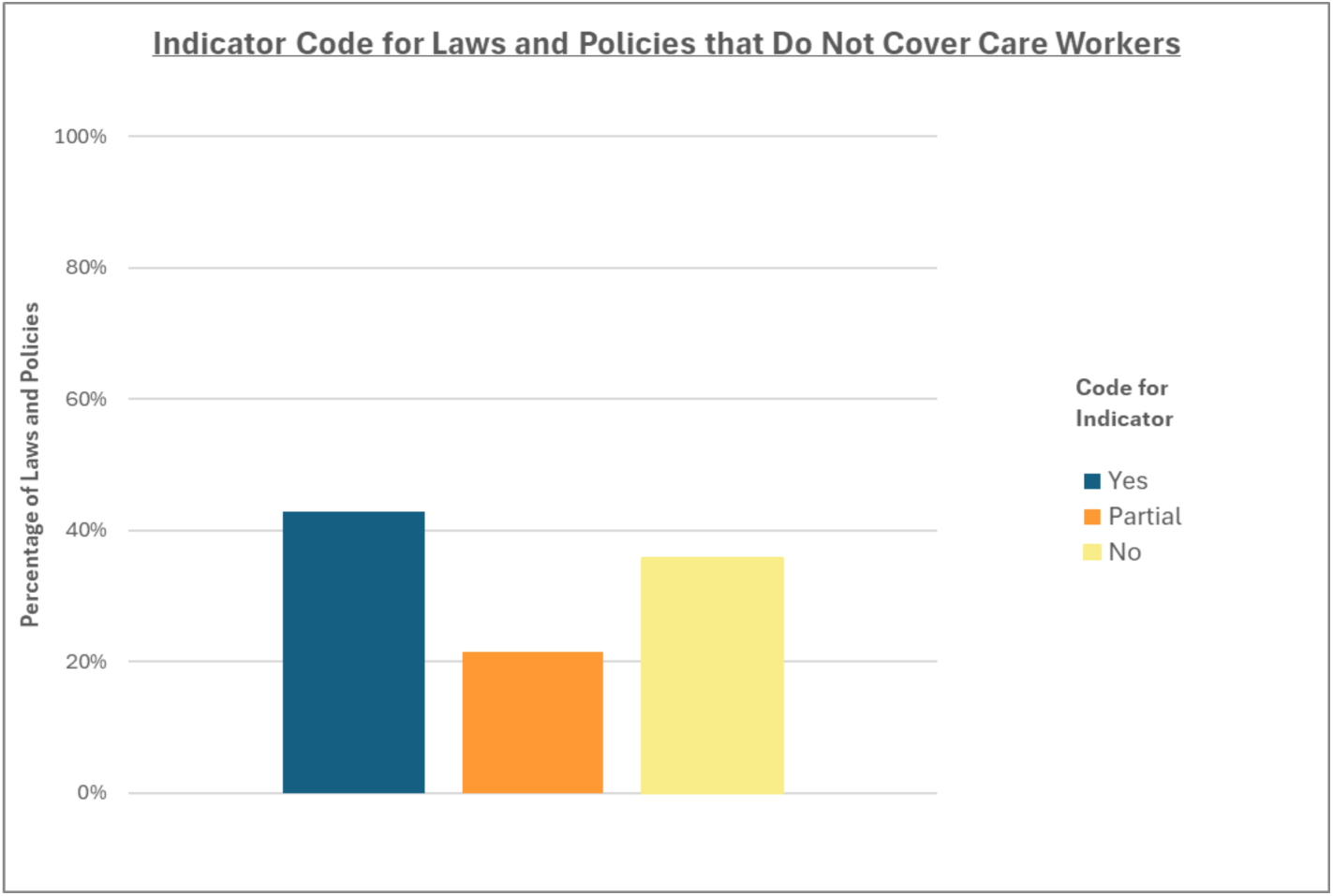
: Alignment of Laws and Policies Not Covering Care Workers With Indicator Criteria.

**Table 3:** Alignment of National Laws and Policies With Care Compact Indicators Across Four Domains.

Occupational Health
| Country | Does the country have national policies or laws that cover care workers that include provision of PPE? | Does the Law include provision of PPE? | Does the law or policy include care workers? |
| --- | --- | --- | --- |
| Angola | Yes | Yes | Yes |
| Argentina | Yes | Yes | Yes |
| Bangladesh | Yes | Yes | Yes |
| Canada | No | No | No |
| Ethiopia | Yes | Yes | Yes |
| Germany | No | No | No |
| Malaysia | Yes | No | Yes |
| Morocco | Yes | No | Yes |
| Turkey | Yes | No | Yes |
| United Kingdom | Yes | Yes | Yes |

Social Protection
| Country | Does the country have national social protection policies or laws that, at minimum, provide unemployment insurance? | Does the law or policy include care workers? |
| --- | --- | --- |
| Morocco | No | No |
| Bangladesh | No | Yes |
| Canada | Yes | No |
| Germany | Yes | Yes |
| United Kingdom | Yes | Yes |
| Malaysia | Yes | No |
| Angola | No | No |
| Argentina | Yes | No |
| Ethiopia | No | No |
| Turkey | Yes | No |

Non-Discrimination Protections
| Country | Does the country have a law that prohibits discrimination in the workplace, on all internationally protected grounds? | Does the country have a law that prohibits discrimination in the workplace, on some internationally protected grounds? | Does the law or policy include care workers? |
| --- | --- | --- | --- |
| Angola | Yes | Yes | Yes |
| Argentina | Yes | Yes | Yes |
| Bangladesh | No | Yes | No |
| Canada | Yes | Yes | No |
| Ethiopia | Yes | Yes | Yes |
| Germany | Yes | Yes | Yes |
| Malaysia | No | No | Yes |
| Morocco | Yes | Yes | Yes |
| Turkey | Yes | Yes | No |
| United Kingdom | Yes | Yes | Yes |

Freedom of Association
| Country | Does the country have national policies or laws that protect the right of workers to join an independent union or similar worker's organization? | Does the law or policy guarantee the right to join a union and ensure the union's independence? | Does the law or policy include care workers? |
| --- | --- | --- | --- |
| Morocco | Yes | No | Yes |
| Bangladesh | Yes | No | Yes |
| Canada | Yes | Yes | Yes |
| Germany | Yes | Yes | Yes |
| United Kingdom | Yes | Yes | Yes |
| Malaysia | Yes | No | No |
| Angola | Yes | Yes | Yes |
| Argentina | Yes | Yes | Yes |
| Ethiopia | Yes | No | Yes |
| Turkey | Yes | No | No |

### Occupational Health

Out of the 10 countries examined, Canada and Germany failed to meet our criteria for occupational health provisions covering care workers. Canada’s labor code regulates federally regulated industries and workplaces but does not include domestic workers or caregivers under its provisions. Germany’s occupational health law neither explicitly includes the provision of PPE nor mentions domestic or informal workers. Turkey, and Malaysia received a partial coding. In Malaysia’s instance, the national occupational health law ensured coverage for care workers but did not explicitly include provision of protective equipment to employees [47]. Turkey’s occupational health law mentions use of PPE but lacked a definitive requirement for employers to supply such equipment to employees [48]. Conversely, five of the countries satisfied the criteria of our indicator and included explicit mandates for PPE provisions to care workers within their national occupational health legislation.

### Non-discrimination

Six of the countries met our indicator criteria for non-discrimination protections for care workers, while four did not. There are no partials. Bangladesh was the only country from our list to have non-discrimination protections under some, but not all, international protected grounds including gender and sex but explicitly excludes care workers [49]. Canada’s national Human Rights Act ensures prevention of discrimination under all protected grounds; however, the Act applies to limited work environments and sectors [50]. Turkey’s labor law also provides protection against international grounds, but only applies to those employed through a formal employment contract, which may exclude some care workers [51].

### Social Protection

Germany and United Kingdom were the only countries that had social protection laws that ensured provision of unemployment insurance to care workers [52-54]. Bangladesh’s labor law ensures care workers are covered, and the law does have severance pay provisions, but it does not guarantee unemployment insurance [49]. Canada, Malaysia, Argentina, and Turkey provide unemployment insurance under their national law but fall short of ensuring care worker coverage [55-58]. Unemployment insurance benefits under national law are available exclusively to employees in the formal sector in all three countries.

### Collective Bargaining

Five of the countries analyzed cover care workers within national laws or policies that uphold the right to collective bargaining, including the right to join a union independent of government oversight. Morocco, Bangladesh, and Ethiopia were coded partial. All three countries have the right to join a union or similar worker’s organizations, but this right was subject to government oversight.[49, 59, 60] Similarly, Malaysia and Turkey have collective bargaining laws that were not completely independent and only applied to formal workers [51, 61, 62].

## Discussion

A notable finding of this review is that the term “care work” or “care worker” was absent from the text of any binding national legal instrument across all 10 countries reviewed, highlighting a significant gap in formal legal recognition of the care workforce as a whole. Some laws did explicitly either exclude or restrict coverage to specific categories of care workers, such as home aides or domestic workers., In particular, we found 3 laws that explicitly excluded certain categories of care work, and 9 laws that restricted coverage to certain sectors. Although our sample size is limited, our findings suggest that while many countries have national laws or policies ensuring occupational health and safety, protection against workplace discrimination, unemployment benefits, and the right to collective bargaining, these rights are often not extended to care workers. Among laws and policies that did not cover all care workers, 6 out of 14 (43%) met the indicator criteria (*Figure Z*). These included laws concerning social protection in Canada, Turkey, Malaysia, and Argentina and non-discrimination protections laws in Canada and Turkey. Among laws and policies that did cover care workers, 19 out of 26 (73%) fully met the indicator criteria being evaluated, 5 out of 26 (19%) partially met the indicator criteria and 2 out of 26 (8%) did not meet the indicator criteria (*Figure Y*).

Laws and policies that did not meet or only partially met the indicator criterion spanned across all WHO regions and every indicator evaluated, demonstrating the need for both more robust policies on rights and protections and for greater care worker coverage globally. Among the 33 laws and policies that partially or fully met the indicator criteria, only 24 (73%) covered health workers. This highlights the need for further progress to ensure care workers receive comprehensive rights and protections that align with Care Compact standards. A prior study that examined laws and policy environments that offer protection to health workers in 182 countries found that 62% of all national laws and policies aligned with all ten indicators assessed - a promising result and close to the 56% we found in our study on care workers. However, our findings may be influenced by the selection of countries with large economies within each region.

Regional variations were not examined due to the insufficient sample size of countries within each region. Nevertheless, we observed that at least one country from every region either fully or partially met the criteria for each indicator. At least one country in each region had laws and policies that ensured care worker coverage. Similarly, at least one country in each of the World Bank income level categories fully with the criteria for each indicator and covers care workers. While our analysis did not find considerable differences between countries across region and income levels, in protecting care worker rights; future research should explore these disparities using a more extensive dataset.

This, a first study of its kind to document laws and policies covering care workers as relates to Care Compact standards, has several limitations. First, our analysis centered on national laws and policies, but state/provincial-level laws could themselves satisfy our coding criteria. For example, some provincial laws in Canada do provide occupational health and protection and cover care workers [63]. Future studies across specific regions, socioeconomic contexts or political structures including federalist systems where provincial law governs some areas could yield insights regarding overall context, as well as knowledge sharing opportunities among countries with similar landscapes.

Second, we analyzed only the legislative requirement, not the practical enforceability or implementation of these provisions, especially with respect to care worker coverage. Furthermore, even where laws did not meet our indicator criteria, countries sometimes addressed the issue through other measures. For instance, Bangladesh’s social protection law lacks unemployment benefits but does provide severance pay. Future research should look at the effectiveness and implementation of laws and policies in covering care workers. The present analysis could be a starting point for future research, in determining where laws are present but not effectively implemented.

Third, this study is limited by our selection of countries with publicly available laws and policies through online databases and government websites, and that are in English or have been translated into English. Fourth, this study is limited by a small sample size. And fifth, our research relied entirely on primary sources. This limitation restricted our study from determining whether these laws were interpreted as to encompass care workers. For example, Bangladesh’s labor act defines a worker as anyone employed in an establishment or industry regardless of whether the terms of employment are expressed or implied [49].

Morocco’s occupational health act applies to any person hired for remuneration, regardless of whether the remuneration is in fact paid [60]. Ethiopia’s law defines worker as “any person who has concluded a contract of employment or any other arrangement with an employer whereby he agrees, directly or indirectly, to perform work for the employer for wages [64]. Our analysis approached these definitions as inclusive of care workers. Future research should include validation through secondary sources and country representatives.

This paper provides critical insight into laws and policies protecting care workers. There exists strong evidence to support the argument that laws and policies shape and impact health and health systems [65]. Policies and laws to protect health and care workforce, inclusive of care workers, has shown to improve health workforce retention rate and satisfaction [66]. Furthermore, benchmarked against the Global Health and Care Worker Compact, findings from this study could provide a foundation for further analysis exploring care worker rights and protections. The upcoming WHO Care Compact full and abridged assessment tools present countries and their stakeholders with further opportunities to conduct a fuller review in all, or some areas covered by the Care Compact. A full understanding of national and subnational protections and safeguards provides a baseline for countries seeking to address health and care workforce challenges like shortages, maldistribution, burnout, departures from service and attracting new workers into the sector.

## Data Availability

Data File is Uploaded.

## References

1. International Labour Organization. Social and labour protection. 2018.

2. Martin CA, Medisauskaite A, Gogoi M, Teece L, Nazareth J, Pan D, et al. Discrimination, feeling undervalued, and health-care workforce attrition: an analysis from the UK-REACH study. The Lancet [British edition]. 2023 Sep 9;402[10405]:845–8.

3. Asha Banerjee, Elise Gould, and Marokey Sawo. Setting higher wages for child care and home health care workers is long overdue. 2021 Nov 18.

4. International Labour Organization. Care work and care jobs for the future of decent work. Geneva, Switzerland: International Labour Organization; 2018.

5. Human Rights Watch. Slow Reform: Protection of Migrant Domestic Workers in Asia and the Middle East. Human Rights Watch; 2010 Apr 27.

6. In Europe, migrant workers fail to receive social benefits despite making contributionsInfoMigrants. 2023 May.

7. World Economic Forum. The Future of the Care Economy. World Economic Forum; 2024 Mar 27.

8. World Health Organization. Fair share for health and care: gender and the undervaluation of health and care work. Geneva: World Health Organization; 2024.

9. Resolution concerning decent work and the care economy, International Labour Organization, 112th Session, Geneva, 2024 Sess [2024]..

10. World Health Organization. The Global Health and Care Worker Compact. World Health Organization; 2023.

11. World Health Organization. Seventy-fifth World Health Assembly. 2022.

12. International Covenant on Economic, Social and Cultural Rights, United Nations, General Assembly Sess [1966]..

13. C187 - Promotional Framework for Occupational Safety and Health Convention, 2006 [No. 187], International Labour Organization, Ninety-fifth Session Sess [2006]..

14. C155 - Occupational Safety and Health Convention, 1981 [No. 155], International Labour Organization, Sixty-seventh Session Sess [1981]..

15. ILO. ILO Declaration on Fundamental Principles and Rights at Work [1998], as amended in 2022. 2022.

16. Convention on the Rights of Persons with Disabilities, United Nations, session 61 of the General Assembly Sess [2006]..

17. Convention on the Elimination of All Forms of Discrimination against Women New York, United Nations, [1979].

18. C102 - Social Security [Minimum Standards] Convention, 1952 [No. 102], International Labour Organization, Thirty-fifth Session Sess [1952]..

19. C100 - Equal Remuneration Convention, 1951 [No. 100], International Labour Organization, Thirty-fourth Session Sess [1951]..

20. C111 - Discrimination [Employment and Occupation] Convention, 1958 [No. 111], , .

21. R202 - Social Protection Floors Recommendation, 2012 [No. 202], International Labour Organization, 101st Session Sess [2012]..

22. International Covenant on Economic, Social and Cultural Rights, International Labour Organization, General Assembly Sess [1966]..

23. C168 - Employment Promotion and Protection against Unemployment Convention, 1988 [No. 168], International Labour Organization, Seventy-fifth Session Sess [1988]..

24. International Covenant on Civil and Political Rights, United Nations, [1966].

25. Right to Organize and Collective Bargaining Convention, 1949 [No. 98], International Labour Organization, Thirty-second Session Sess [1949]..

26. C087 - Freedom of Association and Protection of the Right to Organise Convention, 1948 [No. 87], International Labour Organization, Thirty-first Session Sess [1948]..

27. ILO Declaration on Fundamental Principles and Rights at Work, 1998International Human Rights Law Documents. ; 2018. p. 520–3.

28. Global Jobs Pact, International Labour Organization, [2009].

29. Universal Declaration of Human Rights, United Nations, [1948].

30. ILO 1998 Declaration on Fundamental Principles and Rights at Work, International Labour Organization, Hundredth Session Sess [1998]..

31. Kavanagh MM, Radakrishnan A, Unnikrishnan V, Cometto G, Kane C, Friedman EA, et al. Laws for health and care worker protection and rights: A study of 182 countries. PLOS global public health. 2024;4[12]:e0003767.

32. Liem A, Puspita SS, Fajar, Anggraini L. Securing the rights and health of domestic workers: the importance of ratifying the ILO’s C189. Globalization and health. 2024 Aug 1;20[1]:58–6.

33. ILO global estimates on international migrant workers Third edition ed. Geneva, Switzerland: ILO; 2021.

34. International Labour Organization. COVID-19 and care workers providing home or institution-based care. ILO; 2020 October.

35. Vishakh Unnikrishnan. Who is a Care Worker? O’Neill Institute; .

36. Liem A, Anggraini L, Bariyah, Nasrikah, Lestari E. A long overdue recognition: domestic workers as caregivers for older people in Asia. The Lancet. Healthy longevity. 2023 Apr;4[4]:e129–30.

37. International Labour Organization. International Labour Standards on Freedom of association.

38. Chenwi L. Non-discrimination in socio-economic rights : CESCR general comment. ESR review. 2009 Jul 1;10[2]:27–8.

39. Schoberer D, Osmancevic S, Reiter L, Thonhofer N, Hoedl M. Rapid review and meta-analysis of the effectiveness of personal protective equipment for healthcare workers during the COVID-19 pandemic. Public health in practice. 2022 Dec 1;4:100280.

40. International Labour Organization. C155 - Occupational Safety and Health Convention, 1981 [No. 155].

41. General comment No. 23 [2016] on the right to just and favorable conditions of work, B, , [2016].

42. Asenjo A, Pignatti C. Unemployment insurance schemes around the world evidence and policy options. St. Louis: Federal Reserve Bank of St. Louis; 2019 Jan 1.

43. Conventions, Protocols and Recommendations [Internet]. [cited January 10, 2025].

44. United Nations. International Covenant on Civil and Political Rights.

45. Michaels R. The Functional Method of Comparative Law. In: The Oxford Handbook of Comparative Law. Oxford University Press; 2006.

46. Barak A. Purposive interpretation in law. 1st ed. Princeton [u.a.]: Princeton Univ. Press; 2005.

47. Regulations Under Occupational Safety and Health Act 1994 [Act 514], 1994].

48. Türkiye Occupational Health and Safety Law, Law No. 6331, 2012].

49. The Bangladesh Labour Act, 2006, 2006].

50. . Canadian Human Rights Act, R.S.C., 1985, 1985].

51. Labour Act of Turkey, Law No. 4857, 2003].

52. The Universal Credit Regulations 2013, 2013].

53. Welfare Reform Act 2012, 2012].

54. Social Code - Book III - Employment Promotion, 1997].

55. Unemployment Insurance Law No. 4447, 1999].

56. National Employment Law No.24013, 1991].

57. Employment Insurance System Bill 2017, 2017].

58. Employment Insurance Act, 1996].

59. Labour Proclamation, No. 1156 of 2019, 2019].

60. Law No. 65-99 relating to the Moroccan Labour Code, 2003].

61. Industrial Relations Act 1967, 1967].

62. Act No. 6356 on Trade Unions and Collective Labour Agreements, 2018].

63. Workers Compensation Act, RSBC 2019, 2019].

64. Labour Proclamation, No. 1156 of 2019, 2019].

65. Burns S, Kawachi I, Sarat A. Integrating Law and Social Epidemiology. The Journal of law, medicine & ethics. 2002 Dec;30[4]:510–21.

66. Public Services International. Strengthening the WHO Global Code of Practice on the International Recruitment of Health Personnel: Evidence and recommendations. 2024 Sep 30.

